# Repeated Testing Necessary: Assessing Negative Predictive Value of SARS-CoV-2 qPCR in a Population of Young Adults

**DOI:** 10.1101/2021.03.10.21253292

**Authors:** Elaine E. Thompson, Joseph H. Rosenthal, James Wren, Erik Seetao, Niels H. Olson

## Abstract

Determining when individuals should be released from quarantine is critical for successfully managing a COVID-19 outbreak and local protocols frequently call for testing during the quarantine period, generally after a reasonable incubation period, which raises a question about the interpretation of test results during the quarantine period. We report the negative predictive value of SARS-CoV-2 qPCR tests based on a retrospective longitudinal analysis of 5349 qPCR tests collected from 1227 US service members infected with COVID-19 aboard the USS Theodore Roosevelt (CVN-71) aircraft carrier. In our retrospective evaluation of recovering qPCR-positive quarantined crew members undergoing repeated testing, the negative predictive value is 80% for tests occurring as late as seven weeks following an initial positive qPCR test result. Repeated qPCR testing is necessary to ensure that a once-infected person is no longer shedding viral RNA. When deciding the stringency of exit criteria, we recommend considering local operational and community risk factors.

## Introduction

One of the challenges in managing COVID-19 outbreaks is ensuring that individuals who have been quarantined are not reintroduced to a group of healthy individuals while they are still potentially infectious. Quantitative polymerase chain reaction (qPCR) tests for viral RNA are typically used for demonstrating that COVID-19 infections are resolved because of the high sensitivity and specificity [1]. However, the negative predictive value (NPV) of nasal swab qPCR may be insufficient to determine when an individual is free of SARS-CoV-2 with only one test. Li et al. [2] demonstrated that hospitalized patients with COVID-19 can have mixtures of positive and negative qPCR tests before recovery. Xiao et al. [3] showed a similar pattern in a broader cohort, including some young subjects, but the sample size of 70 was too small to calculate the NPV accurately. Kurcirka et al. explored the change in SARS-CoV-2 false-negative rate of qPCR for subjects over time, showing a 100% probability of a false negative on the first day of infection, and a 67% probability on day four, which the authors note is typically a pre-symptomatic period. At symptom onset, the authors reported the median probability of a false negative reached 38%, and it decreased to 20% by day eight. The authors noted that their study was limited by the heterogeneity of the sample data, which was accumulated from seven previously published studies, and leveraged both oropharyngeal and nasopharyngeal data [4].

We report a retrospective, longitudinal analysis of the value of a negative CDC SARS-CoV-2 qPCR test for quarantined crew members with COVID-19 who are undergoing repeated testing to show evidence of fully cleared infection. Our study consists of 5349 highly specific qPCR tests carried out over the course of COVID-19 infection in a predominantly young, healthy convenience cohort of 1227 active duty service members. In March 2020, the USS Theodore Roosevelt (CVN-71) aircraft carrier arrived in Guam with the beginning of a COVID-19 outbreak.

Shipboard working conditions do not allow service members to follow social distancing guidelines, and thus the majority of the crew was moved off of the carrier and onto the island. qPCR testing of the entire crew was initiated to sequester sick individuals, separate the exposed, and slow the spread of the disease. Symptomatic service members were given testing priority, and those with symptoms of COVID-19 or a positive qPCR test result were isolated in a different location from the rest of the crew. Repeated testing was used to determine when service members had stopped shedding the virus and could return to the ship. The majority of the crew were transferred to quarantine and tested until they were both asymptomatic and had at least two negative qPCR tests in a row, separated by several days. The definitive report of the operation is described in Kasper et. al [5], and the repeated testing from the outbreak provided the large data set for this analysis of the qPCR NPV.

## Methods

Samples for qPCR were collected by nasopharyngeal (NP) swab in viral transport medium. The presence of SARS-CoV-2 infection was determined by rRT-PCR testing with the Thermo Fisher ABI 7500 or the Bio-Rad CFX96 Touch Real-Time PCR Detection System. Samples were processed with either the Qiagen QIAamp Viral RNA Mini Kit or the Roche MagNA Pure 96 instrument for automated nucleic acid extraction, and testing was performed with the Seegene Allplex 2019-nCOV assay test kit or the emergency-use-authorization assay from the Centers for Disease Control and Prevention (CDC) [6]. These assays included the detection of two targets in the nucleocapsid gene, N1 and N2.

Individuals were questioned twice daily for symptoms, and qPCR testing for return out of quarantine was initiated once COVID-19 symptoms had resolved. Some testing of asymptomatic individuals was initiated within the first two weeks of their initial positive test, but most individuals were tested for release from quarantine beginning on day 14, that is, 14 days after their first positive test result. Testing was concluded, and service members were released from quarantine after two consecutive negative qPCR tests.

qPCR test results for a convenience sample of 1227 SARS-CoV-2 qPCR positive individuals were assembled. Crew members who developed COVID-19 before systematic data collection was initiated were not included in the analysis. Testing was performed to meet operational needs, and sailors were tested 1–13 times with a medium number of five tests.Demographics were self-reported and were collected for over 99% of the cohort. To calculate the daily false-negative rates with respect to the course of COVID-19 disease, each individual’s first positive SARS-CoV-2 qPCR test was designated as Day 0. Indeterminate test results were excluded from the analysis. True- and false-negative tests were summed for each day across the testing period and the daily NPV calculated. We assume that SARS-CoV-2 qPCR has 100% specificity and that true re-infection does not occur, therefore once an individual tested positive, any intervening negative test that preceded a subsequent positive test was counted as a false negative. If the final two tests were negative results, they were counted as true negative results. In some cases, often due to indeterminate or rejected results, there was only one final negative result. In these instances, the single negative test was assumed to be true. We ran an alternative analysis with the assumption that single final negatives should be excluded from the data. In that alternative analysis, NPV stayed relatively low (50-60%), but as people recover, the NPV should approach 100%, and thus we rejected that assumption.

## Results

Demographic information for the cohort is shown in Table 1. The majority of the cohort was male and 89.5% were between the ages of 20 and 39. There is broad representation of races and ethnicities, with 4% fewer Caucasians and 5% more African Americans and Hispanic and Latino individuals relative to the 2019 U.S. Census Bureau estimates for the overall U.S. population [7]. In total, 5349 conclusive qPCR tests were performed across the cohort. The NPV for SARS-CoV-2 qPCR gradually increased with time after the initial positive test. Figure 1A shows the daily negative predictive value of the tests performed across the cohort on each day relative to the initial positive test. The negative predictive value was low early in the infection, and it gradually improved during the first two weeks post-diagnosis. However, the NPV remained at or below 80% for seven weeks post-diagnosis on average. Figure 1B shows the total number of tests performed on each day. Tests were infrequent during each sailor’s initial 14 days, after which testing frequency increased to prepare to release individuals from quarantine. The NPV reported in Figure 1A is less reliable on days when fewer tests were performed due to the small sample size. The seven-day moving average on the graph indicates the overall trend. Table 2 lists the average daily NPV for each week post-diagnosis. In week three, when more frequent testing was initiated, the average negative predictive value was only 69%. Table 1 shows the average NPV for each week post-diagnosis. The standard deviation illustrates the broad variability when the total number of negative tests is low. For example, the days with a high NPV in week two were days with very few negative tests, and thus were not reliable. Similarly, while the NPV rises to 91% at the end of the study period, the number of tests is low.

**Table 1.** Demographics

Sex, age, race, and ethnicity. \* Includes race or ethnicity reported by 10 or fewer people.
| **Sex** | **Number** | **Percent** |
| --- | --- | --- |
| Male | 976 | 79.5% |
| Female | 251 | 20.5% |
| **Age** |  |  |
| Under 20 | 48 | 3.9% |
| 20-29 | 776 | 63.4% |
| 30-39 | 320 | 26.1% |
| 40 and over | 80 | 6.5% |
| **Race** |  |  |
| African American | 265 | 21.6% |
| African American, Caucasian | 11 | 0.9% |
| Asian | 60 | 4.9% |
| Caucasian | 725 | 59.1% |
| Caucasian, Native American | 17 | 1.4% |
| Native American | 22 | 1.8% |
| Pacific Islander | 24 | 2.0% |
| Other multiple races * | 32 | 2.6% |
| Declined to answer | 71 | 5.8% |
| **Ethnicity** |  |  |
| American Indian | 19 | 1.5% |
| Filipino | 39 | 3.2% |
| Latin American with Hispanic Descent | 39 | 3.2% |
| Mexican-American | 72 | 5.9% |
| Spanish Descent | 120 | 9.8% |
| Other * | 161 | 13.1% |
| None | 505 | 41.2% |
| Unknown | 272 | 22.2% |

**Table 2.** Weekly average Negative Predictive Value

Average NPV, standard deviation, and number of tests, per week.
|  | Average NPV | stdev | Tests |
| --- | --- | --- | --- |
| **Week 1** | 43% | 24% | 243 |
| **Week 2** | 76% | 20% | 339 |
| **Week 3** | 69% | 9% | 1358 |
| **Week 4** | 78% | 4% | 1079 |
| **Week 5** | 71% | 6% | 823 |
| **Week 6** | 80% | 4% | 656 |
| **Week 7** | 80% | 5% | 432 |
| **Week 8** | 84% | 8% | 289 |
| **Weeks 9 and 10** | 91% | 13% | 125 |

**Figure 1.**
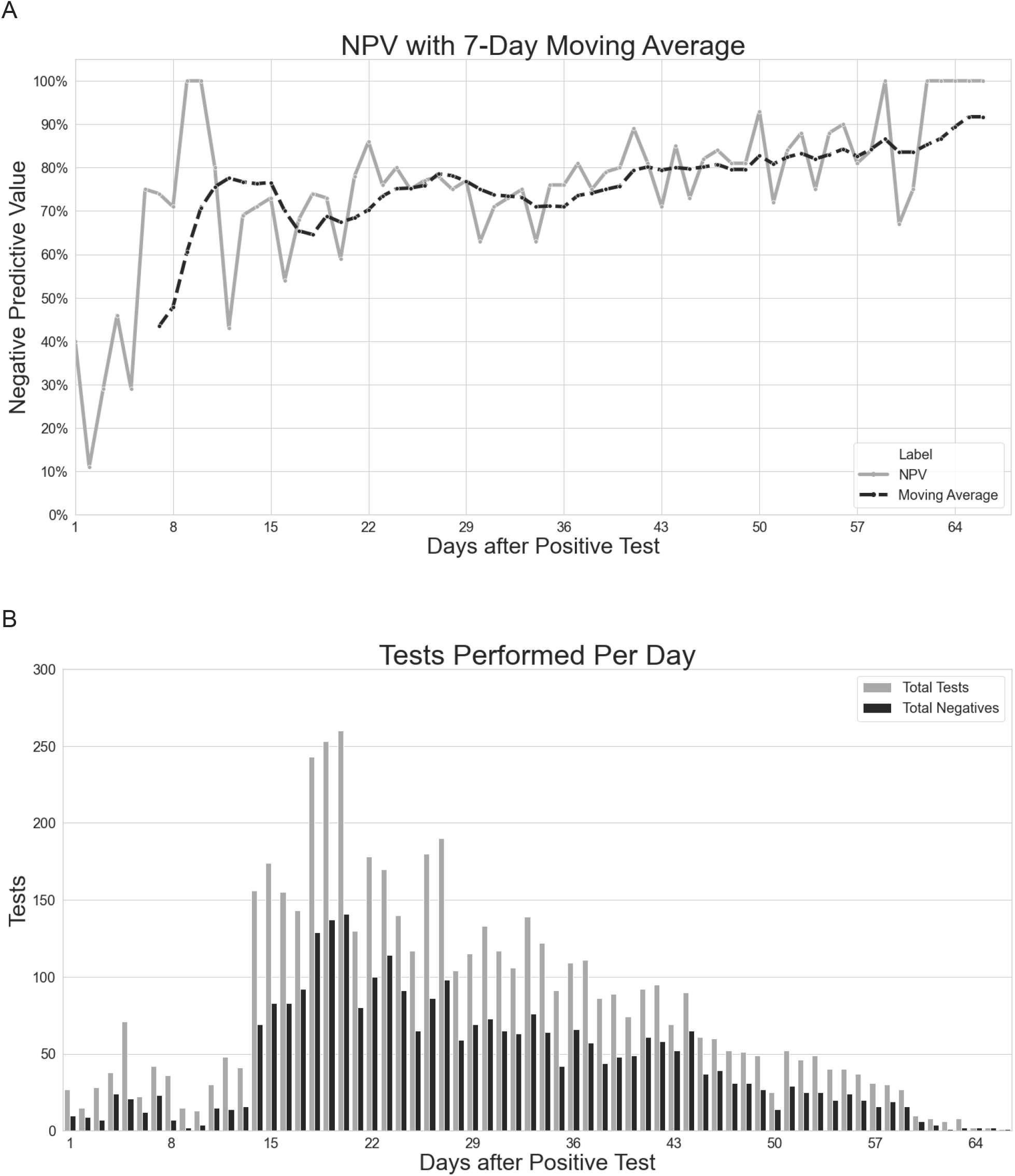
A. Each point indicates the negative predictive value of coronavirus qPCR diagnostic tests performed on that day. The dotted line shows a 7-day moving average of the NPV. B. Total number of definitive tests (gray) and negative tests (black) performed per day.

While the data were assembled to study the NPV of qPCR testing, we noted that many individuals in the cohort remained qPCR positive for extended periods of time. Of the 1227 individuals, 869 or 71% of the cohort, were followed from the time of diagnosis with COVID-19 until they left quarantine after two negative qPCR tests in a row. Figure 2 shows how many people among those 869 had their first negative of the two negative tests in each week post-infection. Strikingly, only 48% had their first of two negative qPCR tests and had presumably cleared the virus by the end of the third week; the remainder continued to have positive qPCR results. 32 people had positive qPCR results until week eight after diagnosis, and six people had positive results until week nine.

**Figure 2.**
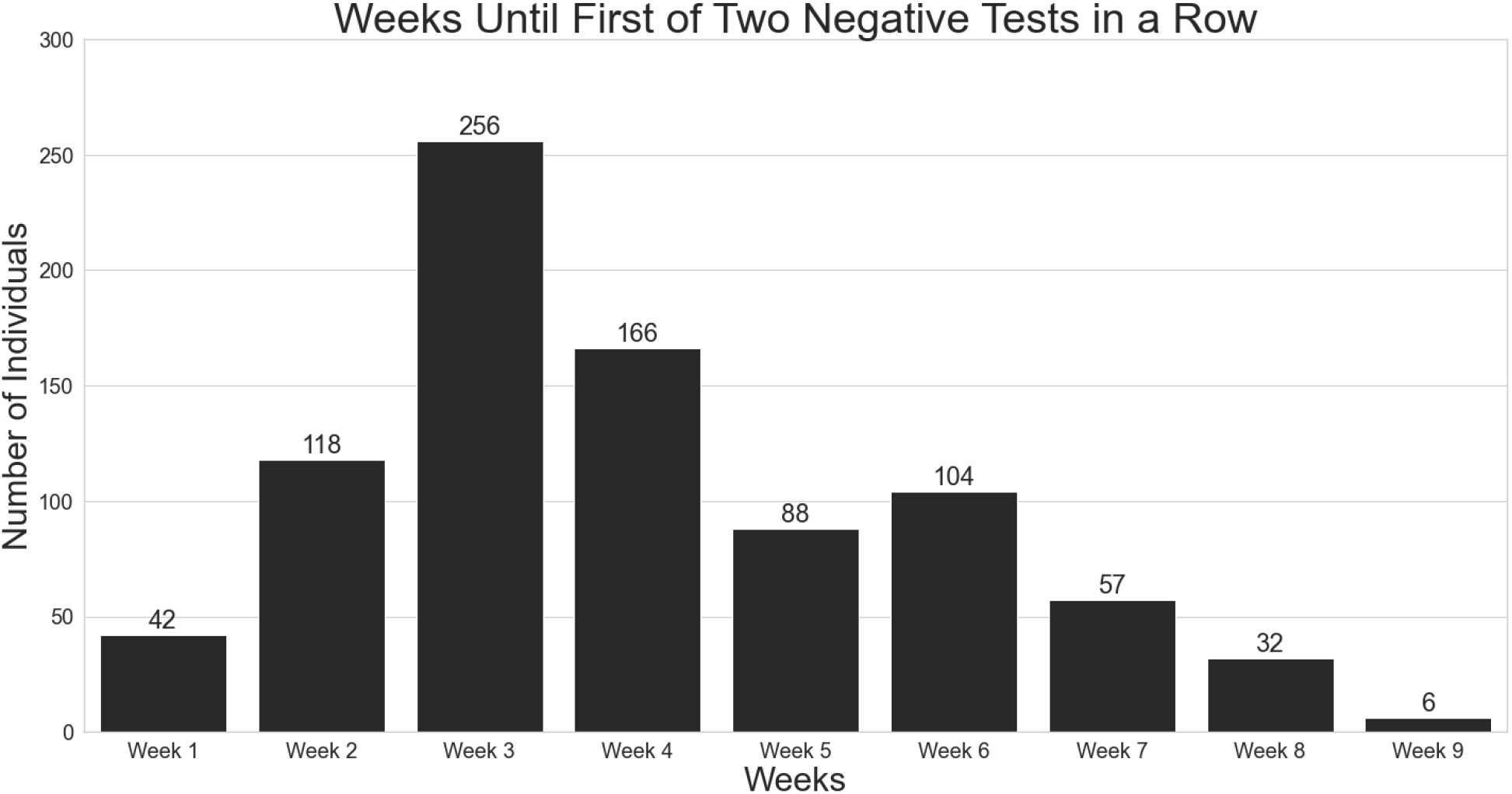
Number of individuals with the first of two negative tests in each week after initial diagnosis. The graph includes 869 individuals. The remaining 358 individuals did not have two negative tests in a row during the period of time available for review.

Our analysis shows a false-negative rate of 20% or higher though week seven. We believed that due to compound probabilities, strings of consecutive false-negative tests could occur Consistent with that hypothesis, we observed 103 cases of two consecutive false-negative test results, 18 cases of three consecutive false-negative test results, and one case each of four and five consecutive negative test results, with each series of false negatives followed by a positive qPCR test. In total 9.94% of the cohort had a string of two or more false-negative tests over the course of their quarantine.

## Discussion

qPCR testing is broadly used both to detect SARS-CoV-2 infection and to clear individuals from quarantine. While a positive qPCR result only indicates the presence of the amplified viral RNA fragment and not necessarily culturable virus, ideally people reentering from quarantine will not have any evidence of potentially contagious SARS-CoV-2. Here, we show that SARS-CoV-2 testing has a low NPV in quarantined individuals, impacting both the ability to detect infections and to determine when a person in quarantine is no longer shedding virus. Additionally, we show that the high false-negative rate can lead to multiple sequential false-negative tests, and that young, healthy individuals can have positive qPCR tests for as long as ten weeks after the initial detection of SARS-CoV-2.

The longitudinal NPV of SARS-CoV-2 qPCR was calculated across 5349 tests collected on a convenience cohort of 1227 young and otherwise healthy U.S. service members who contracted COVID-19. In the first week of infection, over half of the negative tests were followed by another positive test. Sample collection by nasal swab has been demonstrated to be only poorly correlated with viral load in influenza [8], and the false negatives observed in our cohort early in the course of infection suggests that the same may be true for SARS-CoV-2. The NPV is only 70% two weeks post-infection, when many people are feeling better and believe a negative qPCR test means it is safe to leave quarantine. While the NPV improves slightly with time, in a quarantined population, the NPV remains at only 80% seven weeks after initial diagnosis. The high false-negative rate and strings of consecutive tests observed in 9.94% of the cohort suggest that a single negative qPCR test is insufficient evidence on which to base important quarantine decisions. In situations for which the introduction of the virus is exceptionally risky, such as for individuals with key national security roles or those working with extremely vulnerable populations, higher numbers of consecutive negative tests could help to ensure a recovered individual does not exit quarantine while still contagious and informs potential additional manpower requirements in these settings.

The current CDC recommendation is that individuals who were potentially exposed with first degree contacts should be tested initially and a second time 5-7 days later [9]. Not only does the repeated testing mitigate variation in incubation periods, but it lowers the risk of false-negative results from a test with a relatively low NPV. Furthermore, in the subset of this predominantly young and healthy cohort who were tested until they had two negative tests, only 48% had no evidence of virus by nasal swab qPCR for two tests in a row at the end of four weeks, which is twice the usual quarantine recommendation of 14 days. The relatively high false-negative rates among the small number of tests performed early in the course of illness suggest that even initial diagnosis by qPCR should be attempted more than once in populations where prevalence is expected to be high or detection of the virus is critical. Overall, SARS-CoV-2 qPCR has a low enough NPV to necessitate careful interpretation of both tests used for detecting infection and tests used for determining whether individuals are cleared of detectable viral RNA after infection.

## Data Availability

Data available within the article.

## Acknowledgements

Vincent Siu, Naval Information Warfare Command, for organizing qPCR test data.

## Author Declarations

The authors have declared no competing interest.

EET and JHR contributed equally to this manuscript.

The views expressed in this article reflect the results of research conducted by the authors and do not necessarily reflect the official policy or position of The Henry M. Jackson Foundation for the Advancement of Military Medicine, the United States Department of the Navy, the United States Department of Defense, nor the United States Government.

This work was established as exempt by the Naval Medical Center San Diego IRB.

This work was supported by NCRADA-16-471.

I am a military service member or federal/contracted employee of the United States government. This work was prepared as part of my official duties. Title 17 U.S.C. 105 provides that ‘copyright protection under this title is not available for any work of the United States Government.’ Title 17 U.S.C. 101 defines a U.S. Government work as work prepared by a military service member or employee of the U.S. Government as part of that person’s official duties.

